# A Novel Model of Sacral Obliquity, Iliac Obliquity, and Hip Obliquity in Adolescent Idiopathic Scoliosis

**DOI:** 10.1101/2023.08.09.23293754

**Authors:** Pengfei Chi, Junyao Cheng, Kai Song, Bing Wu, Wen Yu, Yang Yu, Geng Cui, Zheng Wang

## Abstract

**Objective:** To propose and validate a novel model to explore the potential compensatory mechanism of sacral obliquity and pelvic obliquity in coronal balance.

**Summary of Background Data:** Sacral obliquity and pelvic obliquity are common phenomena in adolescent idiopathic scoliosis (AIS). However, their compensatory mechanisms in coronal balance remain unclear.

**Methods:** A total of 569 AIS patients were recruited. Coronal pelvic and lower limb parameters, including sacral obliquity (SO), iliac obliquity (IO), and hip obliquity (HO), were measured. Coronal spinal parameters, such as coronal distance (CD), clavicle angle (CA), thoracic curve (T), thoracolumbar/lumbar curve (TL/L), and tilt angle of L2∼L5, were measured. Correlation analysis was performed.

**Results:** The mean values of SO, IO, and HO were 3.34±2.81°, 1.40±2.04°, and 0.82±1.79°, respectively. The largest correlation coefficients with SO were L5 and IO (r=0.725, 0.701). The largest correlation coefficients with IO were SO and HO (r=0.701, 0.801). The largest correlation coefficients with HO were SO and IO (r=0.566, 0.801).

**Conclusion:** SO, IO, and HO have different implications for coronal balance, and their magnitude and direction can be deduced from the novel model. These parameters could be used to evaluate the compensation of the pelvis and lower limbs in AIS. In the coronal position, the pelvis and lower limbs extend the spinal sequence, similar to vertebrae and intervertebral discs.

## Introduction

Spinal deformity is a common clinical manifestation of adolescent idiopathic scoliosis (AIS), often accompanied by sacral obliquity and pelvic obliquity. Whereas how the obliquity of the sacrum and pelvis occurs has not received sufficient attention, and the potential mechanism remains unclear.

When evaluating the posterior-anterior radiographs of patients with scoliosis, we often face the following questions: (1) What is the difference between the commonly used pelvic coronal reference line (PCRL) and the lines connecting the tips of the sacral ala, bilateral iliac crests, and bilateral acetabula? (2) What factors are related to the magnitude and direction of pelvic obliquity? (3) How can we determine whether leg length discrepancy (LLD) is primary or secondary to scoliosis? To answer these questions, we propose a novel model.

## Model

We propose a new model to elucidate the compensation of the pelvis and lower limbs in AIS patients (Figure 1b). The sacrum and ilium are simplified as vertebral bodies, while the sacroiliac joint is a small intervertebral disc, and the lower limbs are a large one, acting as continuations of lumbar vertebral bodies and intervertebral discs.

**Figure 1.**
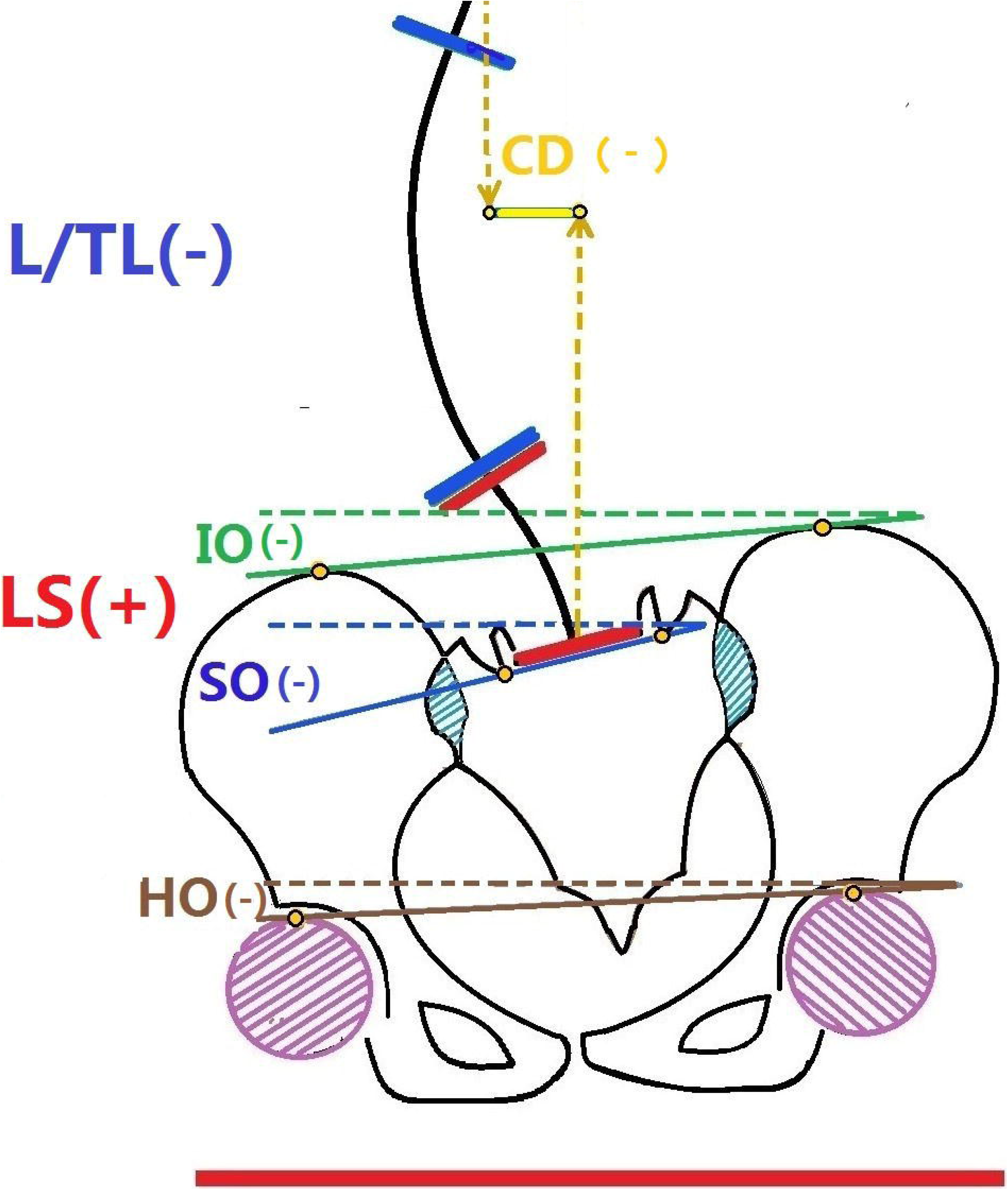
(a) Schematic diagram of spino-pelvic coronal parameters (b) Schematic diagram of the spinopelvic model. CD, Coronal distance; L/TL, lumbar/thoracolumbar curve; LS, lumbosacral curve.

We renamed three angles in this model: (1) Hip obliquity (HO), defined as the angle between the line connecting the apex of the bilateral femoral head and the horizontal line, represents asymmetry of lower limbs, similar to LLD. (2) Iliac obliquity (IO), defined as the angle between the line connecting the bilateral iliac crest and the horizontal line, represents the asymmetry of the ilium and lower limbs. (3) Sacral obliquity (SO), defined as the angle between the line connecting the transition point between the sacral wing and the upper articular process of S1 and the horizontal line, represents the overall asymmetry of the pelvis (sacrum, sacroiliac joint, and ilium) and lower limbs.

The following sections of the study will validate the model.

## Materials and Methods

### 1. Subjects

AIS patients from January 2015 to April 2022 in our center were retrospectively studied. Inclusion criteria: AIS patients with a Cobb angle > 10°. Exclusion criteria: patients with congenital structural abnormalities of the spine, pelvis, and lower limbs; patients with neurological and muscular system abnormalities; patients with lumbar sacralization or sacrum lumbarization.

### 2. Parameters Measurement

Full-length standing radiographs, including the pelvis and bilateral femoral head, were analyzed. Coronal pelvic-lower limb parameters were measured. The parameters were defined as follows: (1) Sacral obliquity (SO), negative [-] if the right edge of S1 was up, positive [+] if left up; (2) Iliac obliquity (IO), negative [-] if the right iliac crest was up, positive [+] if left up; (3) Hip obliquity (HO), negative[-] if the right femoral head was up, positive [+] if left up; (4) Coronal distance (CD), defined as the distance between the C7 plumb line (C7PL) and the central sacral vertical line (CSVL) (negative [-] if C7PL was on the left side of CSVL, positive [+] if on the right side); (5) Clavicle angle (CA): defined as the angle between the line touching the most cephalad aspect of both the right and left clavicles and the horizontal line (negative [-] if the right clavicle was up, positive [+] if left up). (6) Cobb angle of thoracic (T) and thoracolumbar/lumbar curve (TL/L), negative [-] if the convex side was on the left, positive [+] if on the right; (7) Tilt angle of L2∼L5: defined as the angle between the line parallel to the upper endplate of the vertebral body and the horizontal line (negative [-] if the right edge of the vertebral body was up, positive [+] if left up).

### 3. Statistical Analysis

Descriptive statistics were displayed as the mean and standard deviation for continuous variables, and the Pearson correlation coefficient (r) was calculated. SPSS software (version 23, Chicago, IL) was used for statistical analysis. P < 0.05 was considered statistically significant.

## Results

A total of 569 AIS patients were involved in the study, comprising 464 females and 105 males. Regarding AIS types, there were 204 cases of Lenke 1, 85 of Lenke 2, 14 of Lenke 3, 9 of Lenke 4, 244 of Lenke 5, and 13 of Lenke 6. The mean value of each parameter is shown in Table 1.

In Table 2, the correlation coefficient between SO and TL/L was larger than those between SO and the other three parameters. The same relationship also exists in IO and HO. The magnitude and direction of TL/L had an impact on that of SO, IO, and HO, as shown in Figure 2.

**Figure 2.**
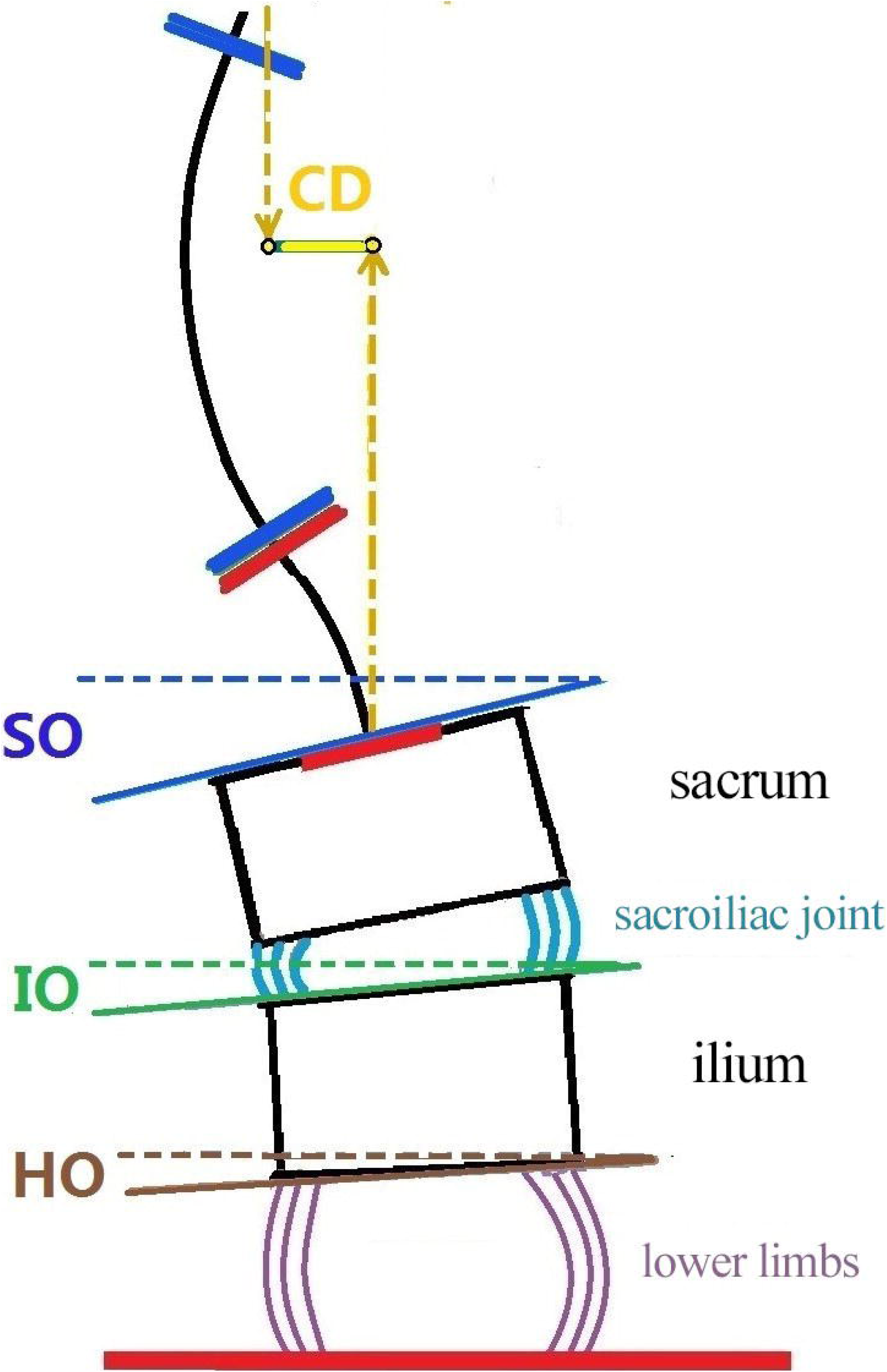
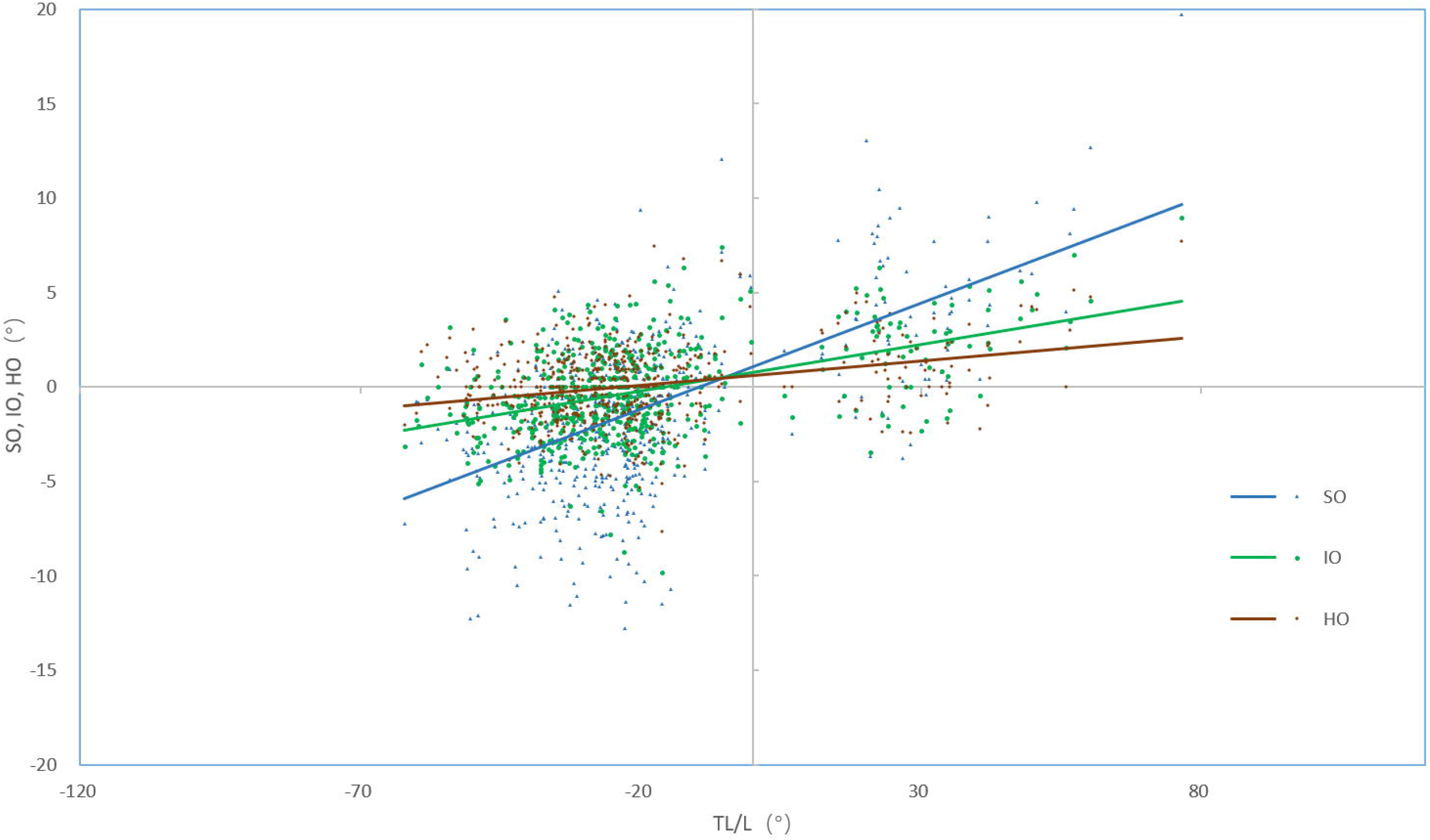
Scatter plot of SO, IO, HO, and TL/L.

In Table 3, we can see that the closer the parameters are, the larger their correlation coefficients will be.

## Discussion

### 1. Significance of coronal pelvic parameters

In 1973, Dubousset et al.^1^ proposed the concept of “pelvic vertebra.” They believed that there were only two non-osseous connections in the pelvic structure composed of the sacrum, ilium, ischium, and pubis—the sacroiliac joint and the symphysis pubis. Since both of the non-osseous connections are micro-movement joints, the entire pelvis can be regarded as an undeformable bony structure after skeletal maturity.

Pelvic parameters are widely used when evaluating sagittal balance. Pelvic incidence (PI) is considered a morphological parameter of the pelvis, which becomes a constant after ontogeny matures. However, a study by Skalli et al.^2^ in 2006 found that in some patients, the change in PI was greater than 5°. In recent years, a series of studies have also proved that PI changes after corrective surgery and during follow-up^3-5^. This indicates that the internal structure of the pelvis is involved in the compensation of sagittal balance, and it also reminds us that the sagittal compensatory ability of the sacroiliac joints and other internal structures of the pelvis are seriously underestimated.

Similarly, based on the concept of “pelvic vertebrae,” previous studies failed to recognize the change of the pelvic internal structure in coronal balance. Therefore, the current definition and measurement of pelvic-lower limb coronal parameters are not unified^6-14^. Several symmetrical anatomical landmarks, including “the tips of the sacral ala,” “the sulcus of the S1 ala,” and “the top of the ilium,” were suggested by the Scoliosis Research Society (SRS) officially to create the pelvic coronal reference line (PCRL). The angle between the PCRL and the horizontal line was defined as the pelvic oblique angle, representing the pelvis’s status in the coronal plane^15^.

To distinguish the different meanings of various parameters and clarify the possible change of the pelvic internal structure in the coronal balance, we renamed the line bilaterally connecting “the sulcus of the S1 ala” as SO, renamed the line bilaterally connecting “the top of the ilium” as IO, and converted the “leg length discrepancy” into angle units and renamed it as HO.

### 2. Significance of the Novel Model

As demonstrated in Table 3, the closer the bony structure is to L4, the greater the correlation coefficient between its angle and the tilt angle of L4. This is also true for L3 and L5. This phenomenon is also observed in SO, IO, and HO. Figure 2 shows that TL/L is linearly related to SO, IO, and HO, with the degree of inclination following the pattern of SO>IO>HO until the distal end touches the horizontal ground. This resembles the distal compensatory curve of spinal deformity, which is increasingly less affected by the primary lesion and becomes smaller until horizontal, both cephalad and caudally. We believe that SO, IO, and HO are fundamentally consistent and continuous with the vertebral body and disc in terms of compensation for coronal balance. As a result, we proposed the hypothetical model shown in Figure 1b.

There are some confusing questions regarding AIS, such as the three mentioned at the beginning of this article. We proposed this model, and its reliability was verified through data analysis. If we treat the pelvis and lower limbs as vertebrae-intervertebral disc structures, the puzzles mentioned earlier can be explained. (1) HO represents asymmetry of lower limbs, similar to LLD to some degree. IO represents asymmetry of the ilium and lower limbs, while SO represents the overall asymmetry of the pelvis (sacrum, sacroiliac joint, and ilium) and lower limbs. The coronal compensatory ability of the pelvis and lower limbs comes from subtle changes in their internal structure, which can be represented by SO, IO, and HO. (2) In patients with scoliosis, it is generally believed that the oblique direction of the pelvis and lower limbs is opposite to the direction of coronal decompensation to ensure overall balance. However, this does not seem to be the actual situation. Our results indicate that the pelvis and lower limbs are not only affected by overall coronal balance but also form part of the local scoliosis sequence. All compensatory curves are centered around the primary lesion, compensating simultaneously in both cranial and caudal directions until they reach a horizontal or nearly horizontal position at the head and feet. For an individual, the factor affecting the magnitude of pelvic obliquity is the distance between the pelvis and the primary lesion. If the main curve is a thoracic curve, the pelvic tilt will be smaller, whereas if it is a thoracolumbar/lumbar curve, the pelvic tilt will be greater. As shown in Figure 2, for SO, HO, and IO, the farther they are from TL/L, the smaller the influence of TL/L and the smaller their angles will be. (3) The relative magnitude of SO, HO, and IO can be used to rapidly determine the source of scoliosis. When spinal curvature is caused by leg length discrepancy, the relative magnitude of the three parameters should be HO>IO>SO, while when the spinal curvature is primary, the relative magnitude of the three parameters should be SO>IO>HO.

This concise hypothetical model allows surgeons to assess the dynamic changes of pelvic parameters in the coronal position and understand the compensatory magnitude and direction of the pelvis and lower limbs more easily. Furthermore, we have used this model to explain some surgical issues, which helps with choosing surgical methods. For instance, the model can predict changes in the compensation of the pelvic and lower limbs after orthopedic surgery. Additionally, we have found that the model can easily solve numerous other scoliosis-related problems, and we plan to conduct detailed research and discussion on these issues in future articles.

### 3. Support for Our Hypothesis from Previous Studies

Schwender et al.^6^ and Lee et al.^7^ confirmed that in AIS patients, sacral and pelvic obliquity mainly occurs when the main curve is located at TL/L segments. Burwell et al.^9,10^ believe that for adolescent girls with lower spine scoliosis, TL/L is related to sacral obliquity, while sacral obliquity is related to lower limb discrepancy. These findings can be explained by our model and data since the TL/L curve is closer to the pelvis, and the sacrum is closer to the lower limbs. Our model also shows the relationship between the thoracic curve and SO, IO, and HO, but the correlation coefficients are smaller because the main thoracic curve affects SO, IO, and HO via the thoracolumbar/lumbar curve.

The study conducted by Cho et al.^8^ analyzed the correlation between sacral obliquity and pelvic obliquity (IO in our study) as well as TL/L curve in AIS patients. The study found a positive correlation between sacral obliquity and both pelvic obliquity and TL/L curve. Additionally, pelvic obliquity was found to be correlated with L4 tilt and LLD. Our model also supports these findings, although Cho’s study did not take into account L5 tilt.

In the study by Pasha et al.^11^, three-dimensional morphological parameters of the pelvis were constructed using the anterior superior iliac spine and posterior superior iliac spine. The study confirmed that 59% of patients with the main thoracic curve had pelvic obliquity towards the convex side of the major curve. Lee et al.^7^ found that the most frequent combination was L4-left type with left-sided sacral slanting. The results of these studies are consistent with our model. However, Pasha’s study found that 79% of patients with the main thoracolumbar/lumbar curve had pelvic obliquity towards the convex side of the main curve, which is contrary to our model. This discrepancy may be due to differences in the selection of measurement points and axial rotation.

Previous studies have shown that there are differences in pelvic bony structure^12,13^ in AIS patients, and the asymmetry of the lower limbs is more of a functional difference rather than an absolute length difference^14^. These findings support our model, which compares the ilium to the vertebra and the lower limbs to the intervertebral disc.

#### 4. Limitations of this study

(1) This is a retrospective study, which mainly conducted the exploration of the general trend and confusing phenomenon. In clinical work, the sacrum (SO) will be horizontal or even incline reversely to the coronal decompensation in some patients with strong compensatory ability in the lumbosacral area (L4/5, L5/S1). This model fails to cover such situations. (2) Patients with lumbar sacralization or sacrum lumbarization were excluded because the anatomical morphology of the junctional area can be irregular, resulting in inconsistent SO measurement. (3) Only AIS patients were enrolled in this study to eliminate the asymmetry of the pelvis and lower limbs caused by non-mechanical factors, thus reducing confounding bias. In theory, the model could apply to other causes of scoliosis.

In conclusion, SO, IO, and HO have different implications for coronal balance. These simple and reliable coronal pelvic-lower limb parameters can be used to evaluate and further analyze the compensation of the pelvis and lower limbs in AIS. Overall, the pelvis and lower limbs extend the spinal sequence in the coronal position, similar to the vertebrae and intervertebral discs.

## Data Availability

All data produced in the present study are available upon reasonable request to the authors.

