## Supplementary material for "A Novel Model of Sacral Obliquity, Iliac Obliquity, and Hip Obliquity in Adolescent Idiopathic Scoliosis": table 1

Table 1 Mean value of each parameter

| Age (year) | 14.67±2.23 (9~20) |
| --- | --- |
| CD (cm) | 0.83±1.71 (-6.02~7.23) |
| CA (°) | 0.65±2.42 (-7.94~8.09) |
| T (°) | -12.32±31.58 (-82.76~70.76) |
| TL/L (°) | 16.15±25.77 (-54.14~76.52) |
| L3 (°) | 6.73±12.31 (-39.25~38.66) |
| L4 (°) | 7.74±10.62 (-24.34~34.93) |
| L5 (°) | 5.51±6.32 (-15.45~32.07) |
| SO (°) | 3.34±2.81 (0~25.59) |
| IO (°) | 1.40±2.04 (-5.61~9.82) |
| HO (°) | 0.82±1.79 (-7.48~7.70) |
