## Supplementary material for "A Novel Model of Sacral Obliquity, Iliac Obliquity, and Hip Obliquity in Adolescent Idiopathic Scoliosis": table 2

Table 2 The correlation coefficients between SO, IO, HO and CD, CA, T, TL/L

|  | CD | CA | T | TL/L |
| --- | --- | --- | --- | --- |
| SO | 0.422^**^ | 0.315^**^ | -0.316^**^ | 0.577^**^ |
| IO | 0.397^**^ | 0.269^**^ | -0.181^**^ | 0.446^**^ |
| HO | 0.100^*^ | 0.176^**^ | -0.231^**^ | 0.290^**^ |

*p<0.05

**p<0.01

CD, Coronal distance; CA, Clavicle angle; T, Cobb angle of thoracic curve; TL/L, Cobb angle of thoracolumbar/lumbar curve
