## Supplementary material for "A Novel Model of Sacral Obliquity, Iliac Obliquity, and Hip Obliquity in Adolescent Idiopathic Scoliosis": table 3

Table 3 The correlation coefficients between parameters.

|  | L3 | L4 | L5 | SO | IO | HO |
| --- | --- | --- | --- | --- | --- | --- |
| L3 | 1 | 0.919^**^ | 0.740^**^ | 0.475^**^ | 0.471^**^ | 0.154^**^ |
| L4 | 0.919^**^ | 1 | 0.871^**^ | 0.637^**^ | 0.548^**^ | 0.251^**^ |
| L5 | 0.740^**^ | 0.871^**^ | 1 | 0.752^**^ | 0.654^**^ | 0.398^**^ |
| SO | 0.475^**^ | 0.637^**^ | 0.752^**^ | 1 | 0.701^**^ | 0.566^**^ |
| IO | 0.471^**^ | 0.548^**^ | 0.654^**^ | 0.701^**^ | 1 | 0.801^**^ |
| HO | 0.154^**^ | 0.251^**^ | 0.398^**^ | 0.566^**^ | 0.801^**^ | 1 |

*p<0.05

**p<0.01
